# Differences in detected viral loads guide use of SARS-CoV-2 antigen-detection assays towards symptomatic college students and children

**DOI:** 10.1101/2021.01.28.21250365

**Authors:** Juan Luis Gomez Marti, Jamie Gribschaw, Melissa McCullough, Abbie Mallon, Jamie Acero, Amy Kinzler, Jamie Godesky, Kelly Heidenrich, Jennifer Iagnemma, Marian Vanek, A William Pasculle, Tung Phan, Alejandro Hoberman, John V Williams, Stephanie Mitchell, Alan Wells

## Abstract

Limitations in timely testing for SARS-CoV-2 drive the need for new approaches in suspected COVID-19 disease. We queried whether viral load (VL) in the upper airways at presentation could improve the management and diagnosis of patients. This study was conducted in a 9 hospital system in Allegheny County, Pennsylvania between March 1-August 31 2020. Viral load was determined by PCR assays for patients presenting to the Emergency Departments (ED), community pediatrics practices and college health service. We found that for the ED patients, VL did not vary substantially between those admitted and not. VL was relatively equivalent across ages, except for the under 25 age groups that tended to present with higher loads. To determine if rapid antigen testing (RAT) could aid diagnosis in certain populations, we compared BD Veritor and Quidel Sofia to SOC PCR-based tests. The antigen assay provided a disease-detection sensitivity of >90% in a selection of 32 positive students and was modeled to have an 80% sensitivity in all positive students. In the outpatient pediatric population, the antigen assay detected 70% of PCR-positives. Extrapolating these findings to viral loads in older hospitalized patients, a minority would be detected by RAT (40%). Higher loads did correlate with death, though the prognostic value was marginal (ROC AUC of only 0.66). VL did not distinguish between those needing mechanical ventilation and routine inpatients. We conclude that VL in upper airways, while not prognostic for disease management, may aid in selecting proper testing methodologies for certain patient populations.

## Introduction

As cases of COVID-19 keep increasing throughout the world and entire populations adapt to living in the context of a global pandemic, accurate rapid testing needs to be available to quickly identify clusters of SARS-CoV-2 infections. Current evidence says that viral load usually peaks in the first week of illness (1) and by the time symptoms develop (2), although patients may not be infectious during the entire process even if viral genetic materials are detected (2, 3). It has been reported that among hospitalized patients, viral load seems to correlate with worse respiratory disease severity, lower lymphocyte counts and increased inflammatory markers, particularly IL-6 and C-reactive protein (4). The strength of association seems to be clearer for plasma viremia, which has been associated with increased risk of mortality (4). However, the utility of plasma virus detection is limited by the fact that only a small fraction of COVID-19 patients presented viremia, with just 44% of those requiring ventilation and only 13% of outpatients found to be positive by upper airway detections of viral genetic materials. Thus, we queried if the viral load in the upper airways can be prognostic for outcome.

The correlation of viral load with clinical course for other common respiratory viruses, such as influenza, is mixed. Some have found an association between clinical symptoms and lymphocyte counts with viral loads, that is not related to age (5). However, the latter study did find that the loads correlated with age, with children having higher levels of respiratory viruses than adults. Similar to this, reports have found significantly larger loads of SARS-CoV-2 among children who are <5 years old (6).

In order to alleviate the demand, cost and time-dependent burden of conducting real time polymerase chain reaction (RT-PCR), more rapid methods of detection have been developed. Rapid antigen tests (RAT) have shown different degrees of sensitivity, and although their sensitivity correlates with viral load levels, some are not advised unless they are used as co-adjuncts of a RT-PCR due to high false negative (FN) rates and overall low sensitivity (7-9). As an example, the Panbio Rapid Ag Test Device, one of the two emergency use list kits of the WHO, has a sensitivity of 11% for detecting positive cases that present with low viral loads, though the assay was found efficacious in detecting those patients that presented with higher viral loads (9). Still, the utility of such tests would depend on whether there is a relationship between viral load and clinical course, or whether some population segments present with higher loads. This impelled us to query the loads across age groups and disease severity to determine utility of these RAT. In short, we find that younger age groups, up through young adults skew towards high viral loads that allow for high probability testing using the RAT. Unfortunately, in the age groups at greater risk for poor outcomes of infection, the lower viral loads negatively impact the utility of such assays.

## Methods

### Ethical statements

All testing was performed as apart of routine clinical care and performed according to CLIA ‘88 regulations by appropriate personnel; all the assays were used under test-specific FDA EUA. The entire study was deemed to be a Quality Improvement initiative by the UPMC IRB and approved by the UPMC QI Review Board; the nucleic acid amplification assays were performed as part of routine medical care, and the antigen testing was added as a quality initiative to determine clinical utilization and interpretation parameters. All patient data were extracted under HIPAA-compliant procedures and de-identified for aggregation.

### Sample collection

Patient specimens for the NAAT testings were acquired by nasopharyngeal (NP) swab placed into viral transport media (VTM). For the antigen/NAAT comparison, dual specimens were collected, one for antigen detection using direct swab testing and one placed in VTM for NAAT. Samples were midturbinate (pediatrics, <18 years) or anterior nares (college-aged, 18-25 years). Antigen tests and corresponding NAAT were administered between March 8 and September 10, 2020. There were five groups of patients. The ED-collected specimens came from nine hospitals in Allegheny County, PA (average population of 1.25 million people). The pediatric specimens were obtaind from symptomatic patients presenting for care to outpatient practices of UPMC Children’s Community Pediatrics and to the Primary Care Center of UPMC Children’s Hospital of Pittsburgh. The students presented to Student Health Services at the Pittsburgh campus of the University of Pittsburgh. The last groups were asymptomatic individuals identified via surveillance testing of either older adults undergoing surgical procedures unrelated to COVID-19 disease, or students at the University of Pittsburgh.

### Nucleic Acid Amplification Test

The NAAT tests used was either Cepheid GeneXpert Xpress SARS-CoV-2 assay (Cepheid; adult inpatients) [Cepheid IFU, https://www.fda.gov/media/136314/download], or Laboratory-Developed Test based on the CDC 2019-nCoV Real-Time RT-PCR Diagnostic Panel EUA (CDC) protocol [CDC EUA IFU, https://www.fda.gov/media/134922/download] (children and adult outpatients). All assays were run per the manufactures’ IFU. Viral loads were approximated based on the C_T_ values obtained from PCR assays, and are presented as such. The C_T_ values for the Cepheid and CDC NAAT testings are not identical but have been shown to be relatively comparable (10).

### Antigen testing

Testing was performed with the BD Veritor and Quidel Sofia2 SARS-CoV-2 assay by testing with the direct swab method, according to the manufacturer’s instructions for use [BD IFU, https://www.fda.gov/media/139755/download][Quidel IFU, https://www.fda.gov/media/137885/download]. The BD Veritor was employed at the University of Pittsburgh Student Health Services for symptomatic students. The Quidel Sofia2 was used in symptomatic children presenting to outpatient pediatric offices. These populations were distinct and non-overlapping with the adult symptomatic patients included in the study, that presented to the Emergency Departments of our nine-hospitals

### Statistical analyses

For sample size purposes, viral loads from LDT and Cepheid platforms were merged for analysis of C_T_ values in Figures 1 and 2. The Kruskal-Wallis test was used to compare more than two groups (Figure 1B, Figure 2A). Pearson’s linear correlation was computed to analyze association between viral load and time period on Figure 1C. For ROC plotting in Figure 2B, the function “geom_ROC()” from “plotROC v2.3.0” R package was used. The AUC was obtained using the model of Mason and Graham (11) with the “roc.area()” function from “verification v1.42” R package. This function calculates the p-value using the Wilcoxon’s test. Figures were plotted using “ggpubr v0.4.0” package, and all analyses and figures were conducted using R v4.0.2.

**Figure 1:**
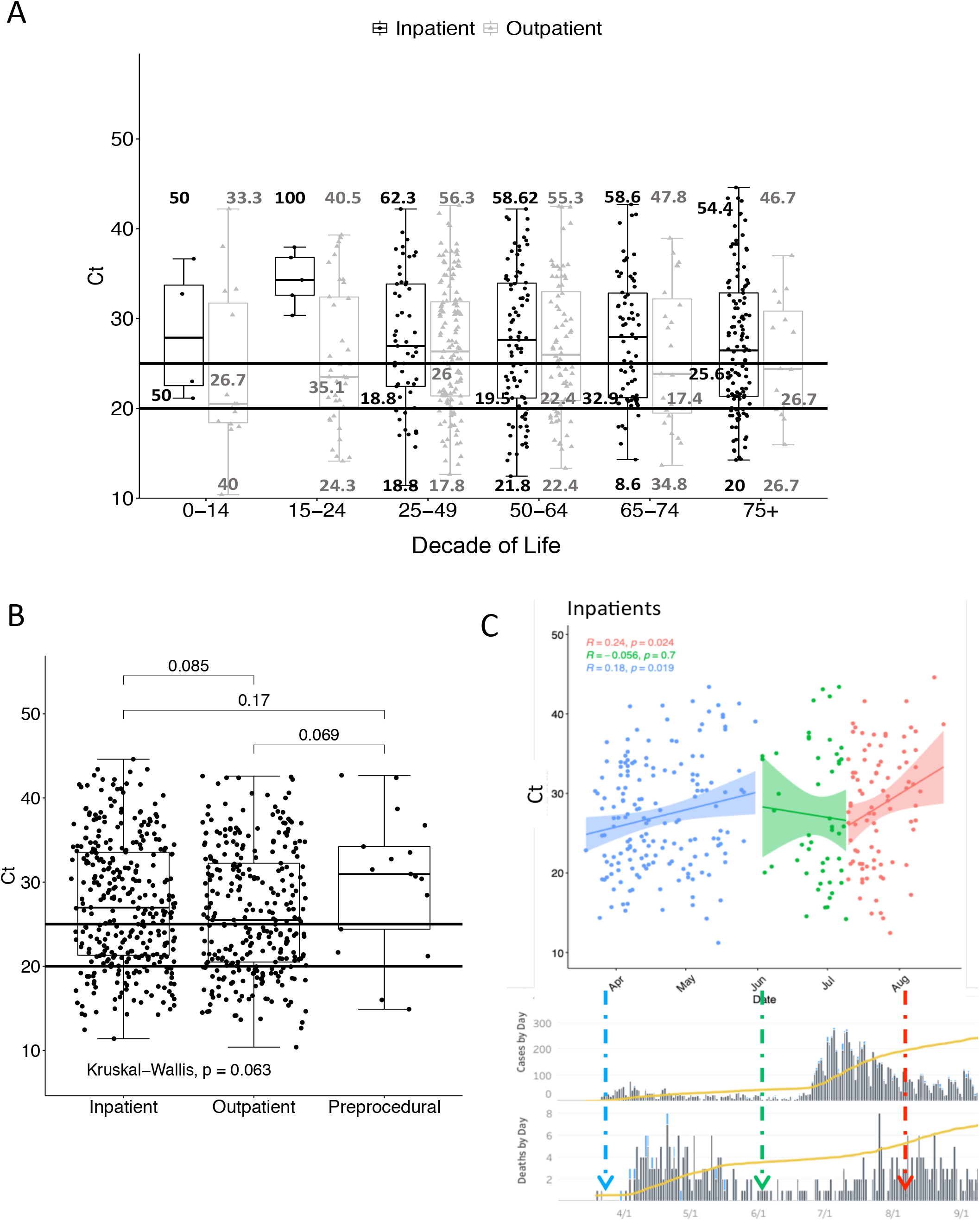
Comparison of viral loads in hospitalized patients in Allegheny county UPMC hospitals. Viral loads were calculated with either the Cepheid GeneXpert or the LDT assay. A) Viral load varies by age grouping in symptomatic inpatients and outpatients. No statistical difference were apparent between the adult populations, but P < 0.05 results comparing the two groups under 25 years old to the older groups. Differences between the inpatient and outpatients are not statistically different (P > 0.05 in each pairwise comparison). B) Viral load appears lower in asymptomatic patients found to carry SARS-CoV-2 upon routine testing prior to undergoing surgical procedures unrelated to COVID-19 disease. C) The viral loads of the inpatients varied with the number of infections found in the resident county. Viral loads trends were detected when the patients were grouped by the level of community infection rate; these were grouped into the initial infection surge (April-May), the interregnum (June) and the later summer surge (August and September), tracked against cases and deaths as compiled by the Allegheny County Health Department (https://www.alleghenycounty.us/Health-Department/Resources/COVID-19/COVID-19.aspx).

**Figure 2:**
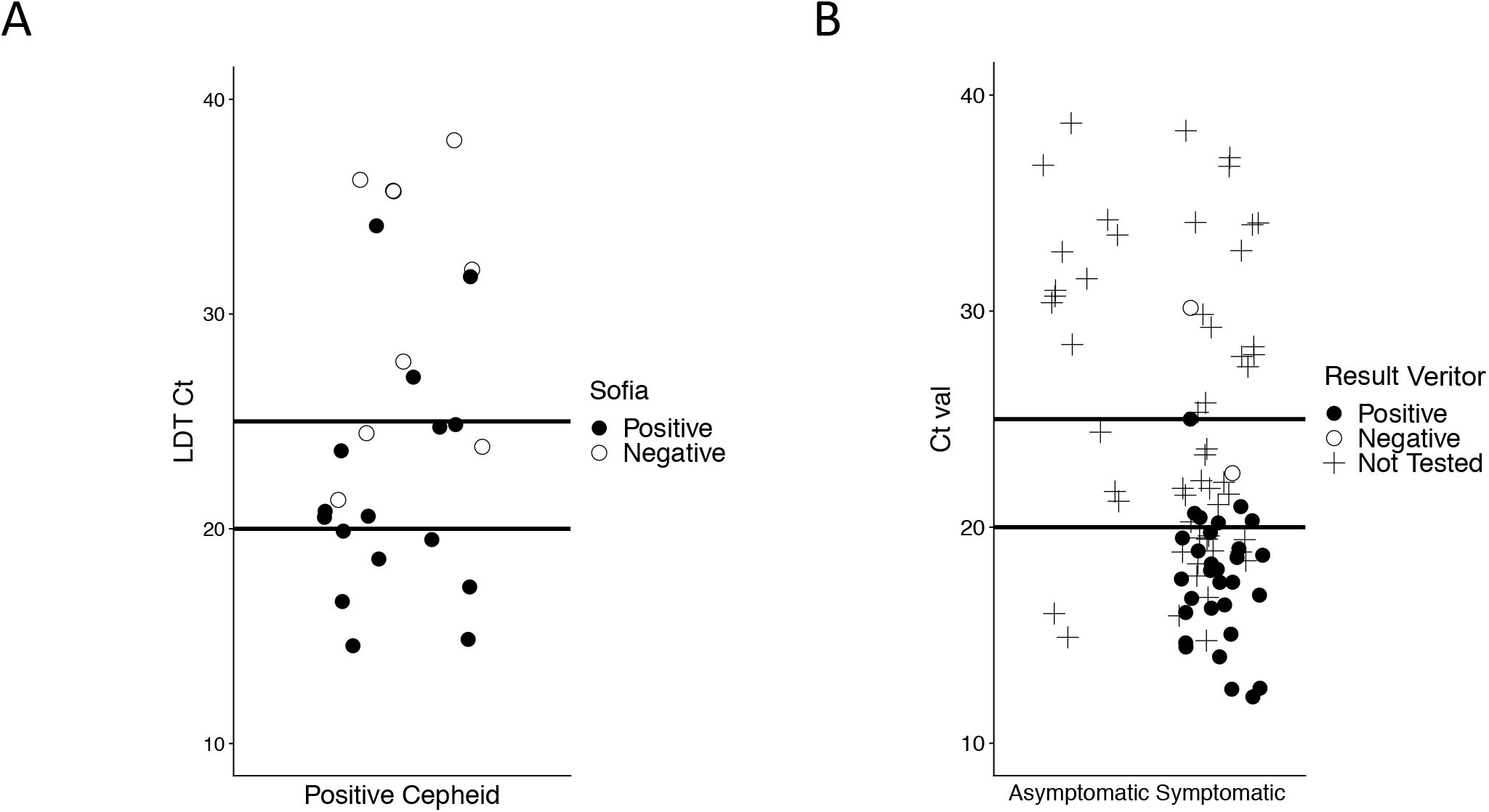
Detection of students and children using rapid antigen testing and nucleic acid amplification assays. Specimens were obtained for both antigen and PCR detection assays. Specimens negative by both methods are not shown. A) Gomez Marti et al. SARS-CoV-2 viral load and antigen testing Children under 18 were tested for SARS-CoV-2 with middle turbinate swabs and tested in parallel using both the Quidel Sofia antigen test and the LDT PCR test. Filled circles represent those specimens detected by the antigen test. B) Students at the University of Pittsburgh were tested by the BD Veritor antigen test and the LDT assay. Asymptomatic students were identified upon random surveillance screening (and not tested with the antigen test), symptomatic students were seen in student health, and subset underwent dual testing. + indicates specimens that were not assessed with the antigen test. Filled circles are those that were positive by the antigen test, and empty circles are specimens that were PCR-positive but antigen-negative.

## Results

### Viral load in the upper airways upon presentation

We tested patients that presented as symptomatic to our acute care facilities. Stratified by age groupings, there was little difference in the adult populations in the perceived viral load as detected by nasopharyngeal swabbing (Figure 1A). However, the detected viral loads in the groups of children (< 15 years old) and young adults 15-24 years old skewed towards higher viral loads (lower C_T_).

Interestingly, the viral loads in persons whose illness did not require hospitalization trended higher (lower C_T_) though these differences did not reach statistical significance in those populations with sufficient numbers for comparisons (Figure 1A). This difference led us to ask whether asymptomatic carriers in our population were similar in viral load. We examined the viral load (mean C_T_ 28.8 ± SD 8.2) in asymptomatic persons (aged 14.6-83.6) who were tested prior to surgical procedures. During this period, we found a carriage rate of approximately 0.25% in this group (in testing of about 30,000 persons across Central and Western PA). The 17 positive adults for which we had C_T_ values tended to have numerically lower viral loads than the symptomatic adults as per higher C_T_ values, although these differences did not reach statistical significance (P = 0.07 compared to outpatient, and P = 0.17 compared to inpatients) (Figure 1B). Viral loads were slightly higher among outpatients compared to inpatients although this did not reach significance (p=0.08).

The viral load in the adults appeared to vary with the prevalence of cases in the community (Figure 1C). Plotting the viral loads against the number of cases in the same county as the hospitals (Allegheny County, PA), we found that higher loads coincided with the initial peak in April and the second summer peak in July.

### Antigen testing in younger outpatients

Based on the higher viral loads in symptomatic patients <25 years old (Figure 1A), we determined whether RAT could be used in these groups (Figure 2). In the pediatric population, the RAT detected 31 of the 43 PCR-positive symptomatic children and adolescents with only 1 antigen-positive assay not being detected by PCR, out of 427 total patients. This resulted in a sensitivity of 72% and a specificity of 99% within a community prevalence of about 10%. In the college-aged population, the BD Veritor antigen test could detect 30 of 32 PCR-positive students. There were no students who were found to be antigen-positive but PCR-negative. This calculates to a sensitivity of 93.75 % and a specificity of 100%; however, this disease sensitivity may represent an overestimate resulting from randomly selected students for Veritor testing included fewer students with lower viral loads (C_T_ > 25) than the students tested only by PCR. If we extrapolate to the total population of symptomatic students, the sensitivity would be about 80% in this group.

### Expected rates of detection by antigen tests

When comparing the RAT positives and negatives against PCR (Figure 2), we find that at viral loads indicated by C_T_ < 20, provided 100% antigen detection by both RAT assays. With C_T_ between 20 and 25, RAT detected most but not all of the PCR positives. At C_T_ > 25, most PCR-positive patients were not detected. We used these C_T_ cutoffs to model expected sensitivity of RAT for symptomatic adults. Modeling for the independent set of patients that presented at the hospital ED, including UPMC Children’s Hospital of Pittsburgh, provided an expected sensitivity of approximately 70%, similar to that found among symptomatic children and adolescents presenting for care to outpatient pediatric clinics. For adults, the expected sensitivities of the antigen tests would be < 50% (Figure 1).

### Viral load in the upper airways and hospital course

Our data suggests that the faster and more available RAT would miss about half of the detected adults, but this would be less important if the detected patients represented the more severely sick patients. To determine if viral load at presentation could be used to manage adult patients in the acute setting, we asked if we could predict the course of hospital stay. The outcomes of adults’ clinical course was stratified based on dying or being ventilated and successfully weaned during the course of that hospitalization. Those who died presented with statistically significantly higher viral loads (Figure 3A) compared those being ventilated or with hospital stays without this intervention. However, patients requiring ventilation did not differ in viral load from those not requiring such. Despite the statistical correlation we found for the group, the viral load at presentation could not be used as predictor of outcome as the Receiver Operating Characteristic (ROC) curve of viral load versus death, had a meager Area Under the Curve (AUC) of just 0.66 (Figure 3B).

**Figure 3:**
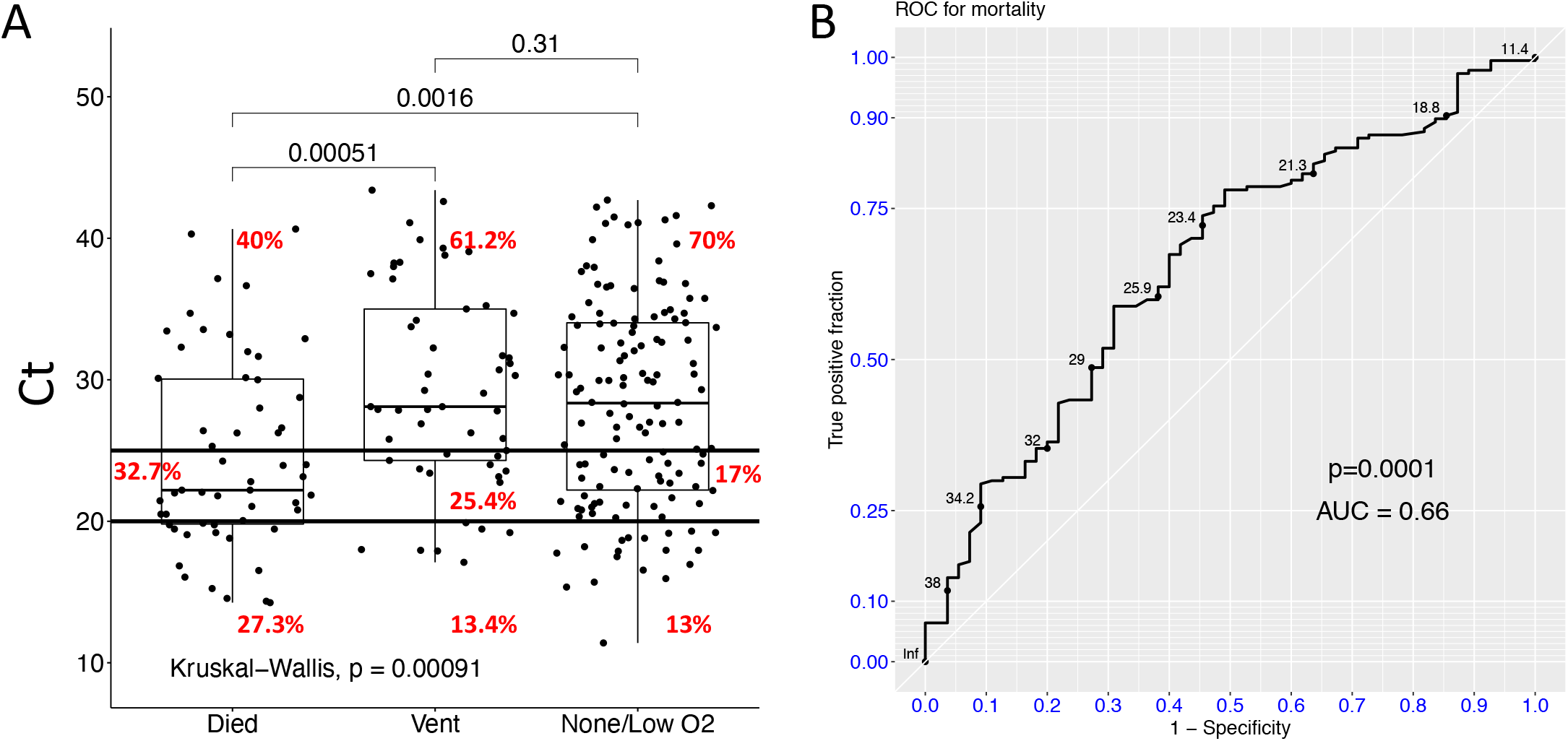
Implications of viral load for outcome of hospitalized patients. A) Viral load is statistically higher in patients who went on to die during the hospital stay when compared to those requiring ventilation or supplemental to no oxygen by mask. However, there was no statistical difference in viral load at presentation between those that required ventilation and those that did not. B) Viral load is a poor marker for death. An ROC, generated to determine whether presenting viral load would designate a mortal outcome, while reaching statistical significance, had minimal power (AUC of only 0.66) to indicate the outcome.

## Discussion

Herein we report on the relationship of viral load, as indicated by CT, upon testing results with clinical outcomes. Our experience included patients who presented to the EDs of the nine hospitals of one health system and were deemed sufficiently symptomatic to be tested for SARS-CoV-2. The study also included asymptomatic patients undergoing pre-procedural testing, and symptomatic outpatient children and college students tested by RAT. Although, these populationss provide a reasonable representation of the US, they still do represent a limited experience.

The focus on viral load at presentation reflects both the ability to test and the implications for selecting test analytical sensitivity. It is well recognized that virus load in the lungs (11) but not in the nasopharynx (4, 12, 13) equates to significant disease in lungs. This connection between viral load in the nasal cavity and disease severity (4, 12, 13) appears to be present in our data with higher viral load correlating with worse outcomes (death) (AUC of 0.66), however with a strength that precludes consideration of causality. Further, the lack of difference between those persons who require in hospital care and those who can be treated safely at home suggests that viral load cannot be used for prognosis or patient disposition.

However, viral load in the nasal cavity has implications for testing approaches. Our data suggests, like other studies, that more rapid and readily available RAT require higher viral loads for detection (14, 15). This requires modeling for proper utilization. Our finding of higher viral reflecting disease density (Figure 1C) would suggest that RAT would perform better for detecting disease during times of infections surges; as such studies that report on high degrees of disease sensitivity of RAT (16) should be interpreted in terms of community prevalence.

One argument for RAT testing in all populations involve some claims that lower viral loads (higher C_T_) reflect carriage that not requiring detection (17). Our data argues against this approach given that lower viral loads (C_T_ > 30) were found at presentation in ∼20% of the patients who died, and ∼30% of those who required mechanical ventilation (Figure 3). Importantly, we found that younger patients tended to present with higher viral loads, a finding that comports with the finding of higher loads of influenza viruses in children (5). As such we tested whether RAT could be used in younger populations (Figure 3). As with RAT for influenza virus, symptomatic children and college-aged persons were detected at high rates. Given limited disease severity in these otherwise healthy young children and young adults, 70% sensitivity may be considered sufficient for use of these point-of-care tests in these groups. However, we still counsel the greater analytic sensitivity of PCR-based testing for high-risk children and young adults, such as those who are transplant recipients or with other underlying comorbidities.

For the older adults, the disease risks and somewhat lower viral loads argues against using RAT to rule out SARS-CoV-2 infection. Modeling suggested that RAT would miss at least half of symptomatic patients who required hospitalization for COVID-19, and even a greater percentage of asymptomatic persons tested prior to a medical procedure. It should be noted that in some cases, such as inclusion in early treatments for pre-hospitalization remdesivir or convalescent plasma/engineered antibodies, the two RAT assays assessed herein, given negligible rates of false positives, might be a reasonable detection modality.

In summary, the viral loads detected in the nasal cavity were found to span broad ranges that do not allow for viral load estimation to guide patient disposition, treatment, or prognosis. Small but significantly higher viral loads in younger persons, coupled with lower disease severity in these persons, suggests RAT may be used as the primary diagnostic modality in these persons, despite the lower analytic sensitivity.

## Data Availability

All data are available as aggregated de-identified data from the corresponding author. All relevant data are included in the manuscript.

## Acknowledgements

This study was enabled by internal funding provided by UPMC Hospital System and the University of Pittsburgh; the funders had no role in study design, data collection and interpretation, or the decision to submit the work for publication.

We thank the many others members of the UPMC Clinical Microbiology Laboratory and Children’s Hospital services and the University of Pittsburgh Student Health Services for helping with specimen collection and testing.

